# An Evolution of Pilot Nutrition Programs to Improve Diet Quality among Older Veterans

**DOI:** 10.64898/2026.09.28.26364174

**Authors:** Jamie Giffuni, Morgan Fique, Sarah Cassatt, Kate Joyce, Linda Krasniewski, Odessa Addison, Elizabeth A. Dennis

## Abstract

Despite the numerous benefits associated with higher dietary quality, many older Veterans do not meet national dietary recommendations. Existing programs are not typically tailored for older Veterans who may experience physical and environmental limitations that hinder their ability to access and prepare healthy foods. This project piloted the incorporation of age-friendly, tailored, virtual nutrition education into an existing health promotion program for older Veterans. Programming followed an iterative design process over two years. Year 1 included 14 weeks of virtual nutrition education on healthy aging paired with weekly produce bags and recipe suggestions. In Year 2, virtual nutrition education shifted to more hands-on learning emphasizing basic meal preparation and adaptive cooking strategies for mobility limitations. Produce distribution in Year 2 shifted to a farmer’s market model, based on feedback from year 1, allowing participants to self-select items. Primary outcomes included program satisfaction, dietary intake assessed via 24-hour recall to calculate the Healthy Eating Index (HEI-2015), and health-related quality of life (PROMIS Global Health). Findings demonstrate the feasibility and acceptability of a virtual nutrition program incorporating age-friendly cooking instruction and adaptive strategies to improve dietary quality among older Veterans.

## Introduction

Older (age ≥65 years) Veterans make up approximately half of the living veteran population.^1^ Older Veterans are more likely to have multiple chronic conditions, be overweight or obese, report a disability (including difficulty with independent living, self-care activities, and walking)^1^, and be sedentary, compared to older non-veterans.^2–4^ Disabilities that contribute to an inability to care for oneself, compounded with existing barriers to healthy aging, increase the likelihood that an older Veteran will require greater Veterans Health Administration (VHA) healthcare utilization. Given that approximately half of the older Veteran population receive care through the VHA,^1^ developing tailored healthy aging programs accessible to these Veterans is paramount.

The Gerofit program is an example of an age-friendly VHA sponsored outpatient exercise and health promotion program targeting older Veterans. Currently offered at 33 VA locations nationwide, as well as virtually, Gerofit provides Veterans with a space to connect with their peers while staying active through personalized exercise plans, guided support, and group classes. Veterans participating in Gerofit have seen improvements in physical and mental function,^5,6^ mobility,^7^ decreased medication utilization and 10-year mortality;^8,9^ however, Gerofit lacks a formal nutrition component within its program offerings. Healthy diets that include fruits and vegetables are linked to a reduced risk of chronic disease including mobility disability,^10^ and are associated with higher muscle mass, strength and physical performance^11–13^, potentially slowing further disability progression later in life. Cross sectional studies demonstrate that healthier dietary patterns, such as those characterized by a higher intake of fruits, vegetables and unsaturated oils are associated with lower rates of disability,^13,14^ than those with less optimal intake.^15^ Longitudinal cohort studies suggest that adherence to a healthier dietary pattern reduces the rate of mobility decline.^15^

Despite the numerous benefits associated with achieving a higher dietary quality, many older Veterans fall short of meeting national dietary recommendations.^16,17^ Our previous work among older Veterans has shown mean Healthy Eating Index (HEI) scores, which assess adherence to national dietary guidelines, were 17% lower than the US national average for adults >65 years.^17^ Fruit, vegetable, and protein intake was also lower in this population as compared to national averages for older adults.^17^ Veterans are likely to overconsume calories from added sugars and solid fats, while falling short of fruit and vegetable recommended intakes compared to non-Veterans.^16^ In addition to established factors that may impact dietary intake and result in poor dietary quality, such as age-related changes in taste, smell and appetite,^18^ older Veterans have been exposed to a military environment where a higher emphasis was placed on maintaining optimal physical performance as opposed to nutrition education.^19^ Mental health, education status, social support, socioeconomic status and food insecurity are also critical factors to consider.^20–22^ This underscores a need for targeted dietary interventions within this group, and mitigating some of these factors could make meaningful differences in diet quality and overall physical health among older Veterans. While the VHA Food and Nutrition Service offers nutrition education through programs such as the Move Weight Management Program (MOVE!) and Healthy Teaching Kitchens, these programs are not tailored for the older adult population, particularly for those who may experience physical and environmental limitations that may hinder their ability to access and prepare healthy foods.^23^ To fill this gap, we developed and piloted the incorporation of an age-friendly, tailored, virtual nutrition education intervention into Gerofit and examined the impact on dietary quality and health-related quality of life among older Veterans.

## Methods

The Gerofit program provides an existing infrastructure into which nutrition programs could be incorporated as a strategy to reach older Veterans across the nation.^24^ Nutrition programming followed an iterative design process, initially implemented in 2021 and subsequently refined based on Veteran feedback for re-implementation in 2022. Participation in nutrition programming was voluntary, and participants did not receive compensation for participation in the classes or assessment completion. The data collected as part of this program was deemed exempt under the University of Maryland, Baltimore Institutional Review Board (IRB #00093722).

### Participant Recruitment

Participants were recruited from the Baltimore VA Medical Center Gerofit program. Gerofit program eligibility criteria has previously been reported,^5^ but briefly, participants must be at least 65 years old, have approval from their VA primary care provider to engage in exercise, and be medically stable, meaning that their existing health conditions are currently being managed by their medical provider and/or other clinicians. For Gerofit participants who wished to participate in either of the nutrition programs, there were no additional exclusion criteria.

### Virtual Nutrition Education Program

In response to COVID-19 pandemic physical distancing requirements required by the VHA, the virtual nutrition education program was developed in 2021. Guided by the social cognitive theory framework, the aging-focused curriculum included 14 modules that focused on self-regulation skills, e.g., goal setting, overcoming barriers and identifying triggers that cause less optimal dietary choices,^25^ as well as general nutrition content relevant to the aging population. Each week focused on a different educational module that included an instructor guide and participant materials. Instructor guides contained instructor notes, key points, and take-away messages providing connected learning week to week. Materials provided to participants included an educational handout, applied activity, and recipes for the produce that was available for weekly pick up (described below). Class materials were emailed to participants, but paper copies were also provided at produce pick up for Veterans who preferred the printed option. Classes were instructed by a registered dietitian with experience in delivering nutrition guidance to Veterans. With progression of the program, additional topics were added based on Veteran feedback and questions, including information about produce preparation, food storage, and basic cooking skills. Planned open discussion time during class allowed for comradery and Veterans to share recipe ideas/suggestions for produce preparation, and additional recipes were shared as requested by participants. Classes occurred weekly for 1 hour over 14 weeks via VA Video Connect (VVC), the VA’s encrypted and HIPAA compliant telehealth platform.

In conjunction with the virtual classes a Produce Program was offered to all participants, free of charge. This program was similar to a community supported agriculture model where members receive regular produce allotments throughout the season.^26^ Through a partnership with a local farm, Veterans were provided with pre-packaged produce pick up bags the day prior to the class that included up to eight different produce items grown locally. Participants did not have control over which items, or the number of items, provided in the bag. An example of a produce pick-up bag included: cucumbers (2), carrots (1 bunch), red tomatoes (2), patty pan squash (2), green beans (1 pint), and green chili peppers (5). Participation in the produce program was optional; participants were not required to take the produce to attend the virtual nutrition education classes. However, program staff recorded the number of participants who picked up their weekly produce. During the Virtual Nutrition program, we were able to ship produce pick-ups bags overnight to a few participants living too far from the Gerofit exercise center to pick them up.

### Virtual Nutrition + Teaching Kitchen Program

In 2022, the nutrition education model shifted to primarily hands-on, experiential learning through an adaptation of the Healthy Teaching Kitchen curriculum. Virtual synchronous group sessions, led by a research assistant under supervision by a registered dietitian, were offered once weekly for four weeks via VVC. The primary reason for the reduction in program length from 14-weeks to 4-weeks was regarding staffing limitations and staff availability. Each session lasted approximately 2 hours, with the first hour focused on educational content and group discussion around a cooking topic, and the second hour included a cooking demonstration that Veterans could follow along with from their kitchen at home. The Healthy Teaching Kitchen modifications included adding aging-related content focused on the Dietary Approaches to Stop Hypertension (DASH) Diet, diabetes, and the importance of protein for older adults, with the incorporation of group discussion. Font size was increased on all handouts for better readability and to accommodate vision changes that may be associated with aging. Based on end-of program feedback from some participants in the Virtual Nutrition Education Program, the lessons focused less on self-regulation skills from the original Healthy Kitchen Curriculum with more emphasis on basic meal preparation and adaptive ways to cook with mobility limitations. Topics that remained the same include an overview of the dietary guidelines, healthy cooking with flavor enhancement through spices and herbs, and understanding food labels. New topics included food safety and storage, setting up kitchens or areas with limited space for cooking, ordering home grocery delivery, and promoting simple recipes, such as maximizing microwave and frozen vegetable use, for older adults on a limited income. Cooking demonstrations included a recipe for breakfast, lunch, dinner, and dessert made with affordable, healthy, easy to find and commonly used ingredients. Veterans were provided with the ingredients needed for the recipes each week prior to the class so they could cook with the group during class demonstrations.

Based on feedback from 2021, the 2022 Produce Program model was transitioned to a farmer’s market style, where Veterans had access to all of the produce items delivered to Gerofit and were able to select the type and quantity of each item that they wanted. They were encouraged, but not required, to take up to eight different types of fresh produce each week. Farmer’s Market items available for pick-up were similar to items offered in 2021 and mostly focused on fresh vegetables grown locally at a farm with proximity to the Baltimore VA Medical Center, for example: yellow squash, zucchini, rainbow chard, cucumbers. Items rotated weekly and available produce offerings were dependent upon the stage of the growing season and farm availability. Similarly to the prior year, participation in this program was optional for those attending the Virtual Nutrition + Teaching Kitchen Program; however, weekly produce pick up was not recorded by staff. Additionally, we were unable to ship produce items to participants living too far from the Gerofit exercise center to pick them up.

### Assessments

Demographic data was compiled from chart reviews and self-reported Gerofit medical history. Height was collected from the electronic health record. Body weight was measured on an electronic scale to the nearest tenth of a pound or obtained from the electronic health record for Veterans who could not make it on-site for their assessment. Height and weight were used to calculate BMI.

Habitual dietary intake (energy, protein, carbohydrate, fat, fruit and vegetable intakes, and fiber intake) was assessed with two 24-hour recalls using the online Automated Self-Administered 24-hour (ASA24) Dietary Assessment Tool, version (2020), developed by the National Cancer Institute, Bethesda, MD, before and after participation, for both programs. Recalls were collected within a two-week period before the program began and within a four-week period after the program ended. The dietary recall was used to calculate diet quality using the Healthy Eating Index (HEI). HEI scores range from 0 to 100; higher scores indicate better adherence to the Dietary Guidelines for Americans.

Participants also completed the PROMIS Global Health Survey. The PROMIS Global Health (v1.2)^27^ consist of 10 items on a 5-pt Likert scale to measure an individual’s physical (GPH) and mental health (GMH). The GPH score comprises 4 items on physical health, physical functioning, pain intensity, and fatigue. The GMH score includes 4 items on overall health-related quality of life, mental health, satisfaction with social activities/relationships, & emotional problems. Raw scores were converted to T-scores for each participant using the T-score look-up tables in the Global Health Scoring Manual.^28^ A higher T-score represents more favorable reports of GPH and GMH.^28^

As an exploratory analysis, we also compared changes in raw scores of individual items of nutrition-related factors, such as pain, fatigue and mental health. Pain was assessed with the question, “How would you rate your pain on average?” Response options ranged from 0 (no pain) to 10 (worst imaginable pain). Fatigue was assessed with the question, “How would you rate your fatigue on average?”, with response options on a 1-5 scale, with 1 being “none” and 5 being “very severe”. Mental health was assessed with the question, “In general, how would you rate your mental health, including your mood and your ability?”, with response options on a 1-5 scale, with 5 being ‘excellent’ and 1 being ‘poor’.

Class attendance was recorded for each program. At the conclusion of each program, participants were asked to complete a short feedback form to assess overall program experience and satisfaction. To assess program acceptability, participants were asked, “On a scale of 1-10 (1 being the worst, 10 being the best), Overall, how would you rate these classes?”. Mean scores were calculated. Participants were also asked open-ended questions regarding what they liked best about the classes, what they didn’t like about the classes, and how the program team could improve the classes. Brief free-text responses were compiled by a program team member and summarized into common themes.

Means and standard deviations were calculated using Microsoft Excel for continuous variables and presented unless otherwise specified, and frequencies were calculated for categorical variables. Paired t-tests were used to assess changes from pre to post assessments.

## Results

Nine Veterans participated in the 2021 Virtual Nutrition Program and completed pre- and post-program dietary assessments. Fourteen Veterans participated in the 2022 Virtual Nutrition + Teaching Kitchen program and completed pre- and post-program dietary assessments. Demographics for each cohort are presented in **Table 1.** Across both groups, the majority of participants were older males with obesity. The Virtual Nutrition Program enrolled a higher percentage of white males, and the Virtual Nutrition + Teaching Kitchen Program enrolled a higher percentage of black males. Veterans attended on average, 11/14 (79%) classes in the Virtual Nutrition program and 2/4 (50%) classes in the Virtual Nutrition + Teaching Kitchen program. Seven Veterans enrolled and participated in both the Virtual Nutrition Program and the Virtual Nutrition + Teaching Kitchen program.

In the Virtual Nutrition Program, 8/9 (89%) of Veterans chose to pick up free produce. On average, those eight people picked up produce on 10/13 available weeks, with the majority of Veterans picking up produce more than 50% of the time. Information on produce pick up from the Virtual Nutrition + Teaching Kitchen program was not kept, as Veterans were free to select any items in any quantity each week.

On average participants expressed a high level of satisfaction for the Virtual Nutrition Program (mean rating: 9.2 out of 10) and the Virtual Nutrition + Teaching Kitchen (mean rating: 9.7 out of 10). Open-ended feedback responses from participants in the Virtual Nutrition Program and Virtual Nutrition + Teaching Kitchen program are presented in **Table 2**, respectively. The open-ended responses were grouped into three common themes: social interaction, produce, and new recipes and information from classes. Despite the generally positive feedback, a few enrolled in the Virtual Nutrition Program expressed concerns about taking the fresh produce given their limited knowledge of food preparation, basic cooking skills, and/or inability to prepare these foods due to physical or environmental limitations (amputated limbs; inadequate kitchen resources and/or space to store produce). This feedback is a large reason why the cooking demonstrations were added to the Virtual Nutrition + Teaching Kitchen program.

Selected dietary intake results and HEI scores are presented by groups in **Tables 3 and 4**, respectively. On average, participants in the Virtual Nutrition program did not improve their HEI score over the 14-week curriculum. Total carbohydrate intake significantly increased in the Virtual Nutrition Program (206±203g vs. 243±243g, p<0.05). There was a trend towards an increase in total fruits consumed per day (1.3±1.0 vs. 1.7±1.4), and a decrease in total vegetables consumed per day (1.6±2.5 vs. 1.2±0.7). There was a significant decrease in daily protein intake in this group by 25 grams (p <0.05). There was no change in energy, total fat, or fiber intakes. However, when examining changes in dietary quality among Veterans who received >80% of produce bag pick-ups (N=5), HEI score increased by 5.4 points and daily fruit and vegetable servings consumed increased by 1.9 servings (data not shown).

On average, participants in the Virtual Nutrition + Teaching Kitchen program improved their HEI score over the 4 weeks of classes. There was no change in energy intake (1943±874 vs. 1970±859 kcal), total carbohydrate intake (185±98g vs. 157±100g), total fat intake (70.8±40.3g vs. 60.3±43.6g), fiber (13.8±5.2g vs 14.7±6.7g), fruit servings (1.0±1.2 vs. 0.8±0.7 or total protein intake (63.5±29.8 g vs. 67.5±32.2 g). Servings of vegetables trended towards increasing over this program (1.2±0.9 vs. 1.6±0.6).

Six participants in the Virtual Nutrition Program completed the PROMIS Global Health questionnaire at baseline and post. T-scores for the PROMIS global physical health (GPH) (48 vs. 47) and mental health (GMH) (48 vs. 48) subscales did not change from pre to post-program; however, were both lower compared to the general US reference population T-score of 50.^28^ On average, participants rated their mental health as “good” at the start of the program and “very good” at the conclusion. Feelings of fatigue (baseline: 3.5±0.5 vs post: 3.6±0.9) and 7-day average pain scores (baseline: 2.5±1.9 vs post: 3.6±1.5) did not change.

Thirteen out of 14 Virtual Nutrition + Teaching Kitchen participants completed the PROMIS Global Health questionnaire. T-scores for the GPH (42 vs. 45) and the GMH (47 vs. 49) improved over the program towards the general US reference population T-score of 50.^28^ On average, participants viewed their mental health at baseline as “good” and this did not change over the program. Feelings of fatigue trended towards improvement from baseline to post-program (2.9±1.1 vs. 3.6±0.9). There were no changes in pain scores (4.6±2.9 vs. 3.9±2.5).

## Discussion

The data collected from these programs demonstrate the feasibility and acceptability of administering virtual nutrition programming that incorporates age-friendly virtual cooking classes as a strategy to improve diet quality among older Veterans. Our prior data suggests that dietary quality, and fruit, vegetable, and protein intake is lower in this population as compared to national averages for older adults,^17^ further justifying the need for tailored programs to improve these outcomes. Among older adults, impaired mobility, lack of cooking skills, and changes in social support are known barriers to consuming a healthy diet. Further, in Baltimore, nearly 25% of older adults live in Healthy Food Priority Areas (HFPA)^29^, urban areas where availability of unhealthy foods exceeds healthy foods which limits accessibility of healthy foods. Older Veterans may have additional challenges in acquiring a healthy diet. Following military service, it is known that physical activity is significantly reduced, but a high-calorie, low-nutrient diet may be maintained in certain individuals.^9^ As the population of older adults, including older Veterans, continues to grow, interventions to improve health and maintain functional independence are more important than ever.

A previous pilot study by our research group showed that participation in an in-person group nutritional education program curriculum resulted in significant increases in self-reported perceived dietary quality and self-reported vegetable and fruit intake among older Veterans.^30^ In our Virtual Nutrition education program, we did not observe overall changes in dietary quality, but we saw a decrease in protein. This prompted further emphasis on protein intake in our Virtual Nutrition + Teaching Kitchen program. Additionally, the open-ended feedback provided insight into barriers to utilizing fresh vegetables, which may have been a potential reason for why vegetable intake may have decreased over the 12-week program. Overall mean HEI improved with the Virtual Nutrition + Teaching Kitchen program, indicating that our intervention did change dietary habits despite the short 4-week timeframe. There was a non-significant trend towards an increase in vegetable intake. This may be because although there was a substantial change in the positive direction, there was a wide standard deviation, which indicates the program may benefit some more than others. It will be important to better understand the factors that determine why some participants may benefit more than others. It will be of interest to investigate how future interventions can further improve vegetable, protein and overall dietary quality, more specifically to prevent loss of lean mass commonly associated with aging. Additionally, 4-weeks is a relatively short time frame, future studies need to explore the impact of a longer intervention.

We acknowledge the relatively short-term impact on dietary quality. However, data suggests that improving dietary intake may positively impact functional outcomes and management of other chronic diseases. Despite our programs showing no impact on pain and fatigue scores, prior literature suggests that diet improvements occurring in middle age were associated with better physical functioning in older age,^31^ and modest changes in diet quality were associated with positive changes in self-perceived physical functioning.^32^ Diet quality is negatively associated with perceived fatiguability^33^ and body pain,^34^ which could impact future functional status. Our program only observed changes in fatigue after the Virtual Nutrition + Teaching Kitchen program. Future work with larger sample sizes and use of validated must explore how these nutrition-related factors may be associated with diet quality and functional outcomes.

The feedback, including the open-ended responses from participants, indicates high satisfaction with the program, especially with the produce and groceries supplies given and the social interaction conferred by the program. The iterative process that allowed for incorporation of Veteran feedback into our programming, and modifications to enhance utilization of our programs ensures that our program is tailored for this population to consider cultural adaptations. We solicited suggestions from Veterans on how to consider the differences in food preferences, accommodations for disabilities that may limit cooking preparation as well as varying cooking skill abilities to better focus on recipes that are simple and familiar/preferable. At the same time, Veterans had limited knowledge of how to utilize certain produce that may be similar to items they were familiar with, and we were able to help them expand and try new produce through our efforts. For example, Veterans didn’t realize that dark leafy greens like kale could be cooked similarly to collard greens. In our programs, Veterans expressed to our staff that they lacked knowledge of how to prepare unfamiliar produce items and that they needed additional help with that. We believe one of the major strengths of this intervention is not only our tailored intervention, but the inclusion of the group discussion that allows participants to discuss how they are able to use the produce for themselves and use this as an opportunity to teach their peers as well as teach the study team. Future interventions may consider extending group discussion time and including more fruit and protein-based food sources in the grocery supplies to help mitigate negative changes in dietary outcomes.

Our programming demonstrated several strengths, including the innovative use of virtual group classes for an intervention for a population often excluded from research due to transportation issues, mobility concerns, and multiple comorbidities. The virtual nature of the classes also allowed for the ability of rural Veterans living outside metropolitan Baltimore to participate. Despite this program being offered solely to Veterans, Veterans who use Veterans Affairs facilities are more likely to be single, with lower levels of education and income than those who do not utilize Veterans Affairs healthcare services.^35^ They also have more comorbid illness and obesity rates than those who receive care from non-Veterans Affairs facilities,^2^ which suggests interventions to improve overall health are warranted in this population. Successful implementation of an intervention to meet the needs of this population will likely ensure that we address barriers that are also prevalent among the general population; thus, the intervention materials could easily be implemented and disseminated to non-Veteran populations.

Our programming also encountered some limitations. First, habitual dietary intake was self-reported using the average of two weekday 24-hour recalls before and after the programs. The weekday status may have influenced both HEI scores before and after the intervention to be slightly higher than expected.^36^ While there is data to suggest fewer variations across dietary intake between weekend and weekdays in older adults,^37^ additional recalls should be included in future evaluations. Another limitation is the smaller sample size consisting mostly of males living within proximity to the Baltimore VA facility. Future work may need to identify strategies to further extend and advertise nutrition resources to older female Veterans as well as those who live farther away. Strategies to increase engagement for these individuals should incorporate ways to deliver produce and materials to participants’ homes regardless of location. We acknowledge the limited use of the open-ended feedback surveys to assess acceptability. Future studies should include more detailed qualitative assessments such as semi-structured interviews and consider a priori analysis to ensure rigorous methods are employed.^38^ Lastly, the programs only included older Veterans enrolled in Gerofit who also self-selected to participate. As Gerofit is an exercise program that emphasizes health and wellness, those who are enrolled in Gerofit may have certain attributes such as high levels of health mindfulness that may not be representative of the older general VA population. Furthermore, no information was collected on veterans who opted out of the program.

In conclusion, our program demonstrates a feasible strategy for tailoring and implementing an age-friendly virtual nutrition education program for older Veterans. With a better understanding of the nutritional teaching needs of our older Veterans, we hope to be able to implement nutritional education as a long-term program that complements Gerofit in the future.

## Biographical Notes

- **Jamie Giffuni** is a research-trained exercise physiologist who has been employed at the Baltimore VA Geriatric Research Education and Clinical Center for the past 15 years. She works on both the clinical exercise rehabilitation program, Gerofit, and research projects that focus on the performance of exercise interventions in older adults with multiple chronic conditions. The overall hypothesis of these research studies is that there are declines in functional performance, cognition, and metabolic profiles that occur as a result of primary (biologic), secondary (lifestyle), and tertiary (disease) aging which can be attenuated by exercise and lifestyle interventions. Her role in the research is as a clinical research coordinator, performing all aspects of research implementation and assisting with protocol design.
- **Morgan Fique** is a qualitative research analyst in the Department of Physical Therapy and Rehabilitation Science at the University of Maryland School of Medicine. She earned her bachelor’s degree in Exercise Science from Towson University and recently completed the STAR-PREP Postbaccalaureate Program at the Marlene & Stewart Greenebaum Comprehensive Cancer Center at the University of Maryland School of Medicine. Her work focuses primarily on research involving older adults and older Veterans. Morgan’s research interests include using community-based participatory approaches to address social determinants of health and identify preventive strategies that promote successful aging while reducing health disparities among aging populations.
- **Sarah Cassatt, MS, RDN** is a Health System Specialist at the VA Maryland Health Care System (VAMHCS). She previously held the positions of Registered Dietitian, Advanced Practice Nutrition Specialist, and Associate Chief of Clinical Nutrition at the VAMHCS. She is also a doctoral candidate in Gerontology at the University of Maryland, Baltimore where she has conducted research with the VAMHCS, Geriatric Research Education and Clinical Center (GRECC). She received both her bachelor’s and master’s degrees in nutrition from The University of Pittsburgh and Case Western Reserve, respectively. She completed her dietetic internship at the Louis Stokes Cleveland VA Medical Center. Her research as a graduate student has focused on diet, health outcomes, and malnutrition prevalence and risk factors in older Veterans.
- **Kate Joyce, MBA** is the Executive Director of DBA Serviceberry Farm (formerly TALMAR), an organization that provides therapeutic programs focused on horticulture and agriculture to foster vocational training and skill development particularly for individuals with disabilities or chronic conditions that impact their health and well-being. She completed her MBA from the University of Maryland - Robert H. Smith School of Business and BA from Syracuse University.
- **Linda Krasniewski, MD**, is an internal medicine resident at the University of Maryland Medical Center. She received her bachelor’s degree in Molecular Biology from Johns Hopkins University and her medical degree from University of Maryland School of Medicine. Prior to starting medical school, she spent two years at the National Institute on Aging conducting research on sarcopenia in aging. While in medical school, she conducted research at the Baltimore VA Medical Center studying the effect of nutrition interventions in veterans. She plans to work as a primary care physician upon graduation from residency.
- **Odessa Addison**, **PT, DPT, PhD**, is an Associate Professor in Physical Therapy and Rehabilitation Science at the University of Maryland School of Medicine, and a Research Health Scientist in the Maryland VA health care system. Dr. Addison’s work centers on myosteatosis, balance, mobility, and fall risk in older adults, examining how lifestyle factors such as exercise and diet influence muscle health and functional outcomes. She has authored numerous peer-reviewed publications and presents nationally and internationally on rehabilitation strategies to optimize function and independence in older adults.
- **Elizabeth Dennis, PhD, RD** is an Associate Professor in the Department of Physical Therapy and Rehabilitation Science at the University of Maryland School of Medicine, and member of the University of Maryland Greenebaum Comprehensive Cancer Center. She also has a Without Compensation (WOC) appointment at the VA Maryland Health Care System through the Geriatric Research Education and Clinical Center (GRECC). She received both her bachelor’s degree and PhD in Human Nutrition, Foods and Exercise from Virginia Tech, and completed her dietetic internship at Marywood University. Her research at Virginia Tech focused on beverage intake and adult weight management, and during her post-doctoral training, she explored dietary intake habits of African American breast cancer survivors. At UMSOM, her work focuses on lifestyle interventions to improve health and chronic disease-related outcomes, particularly in older adults with multiple comorbid conditions.

## Author Contributions

JG and EAD were responsible for data collection, data management, analysis and/or interpretation of data and drafting the manuscript. LK and SC were responsible for data collection and contributed to manuscript development and revisions. JG, MF, SC, KJ, LK, OA and EAD advised on all aspects of the manuscript and contributed to manuscript revisions. All authors have read and agreed to the published version of the manuscript.

## Data Availability Statement

Supporting deidentified data may be available to bona fide researchers upon request, subject to VA approval.

## Acknowledgements

We would like to thank the Gerofit team, including staff exercise physiologists and other VA staff who assisted with the program, and the Veterans who participated in our programs. We would like to thank DBA Serviceberry Farm staff for providing the produce. Hormel Foods donated sauces and coupons for Veterans participating in the nutrition classes.

